# Adverse Weather Conditions can have Negative Effects on Birth Outcomes: Evidence from a birth registry cohort in Tanzania

**DOI:** 10.1101/2024.01.24.24301731

**Authors:** Rita T. Amiel Castro, Beatrice Marti, Blandina T. Mmbaga, Tobias Siegfried, Thomas Bernauer, Ulrike Ehlert

## Abstract

Climate change is bound to have particularly serious implications for public health in the least developed countries. Based on unique registry data from the Kilimanjaro Christian Medical Centre in Tanzania from 2001-2015, we aimed to investigate whether pregnancy exposure to weather conditions affects neonatal birthweight, length, head circumference, and Apgar scores and to evaluate changes in weather conditions across the studied period. Methods: N=30,068 pregnant women were assessed through a postpartum interview whereas baby data were obtained through medical records. Environmental data - rainfall, temperature, the multivariate ENSO index (MEI), the effective drought index, and harvest - were obtained through the Kilimanjaro International Airport weather station and examined during pregnancy and a preconception trimester. Our main analytical strategy was hierarchical regression analyses. Results: Analyses revealed a median birthweight of 3,185 g (IQR:600), a mean length of 49 cm (IQR:71) and a mean head circumference of 34 cm (IQR:24). Lower birthweight was associated with consecutive days with temperatures >30°C (*B* -.01, CI95% -.00 - .05) in the third gestational trimester, poor harvest (*B* -.13, CI95% -.10 -.08) and the interaction between insufficient rain and poor harvest (*B* .20, CI95% .13 -.26). Shorter length was significantly associated with more days with temperatures >30°C at preconception (*B* -.01, CI95% -.00 - .04) and in all gestational trimesters (range: *B* -.01 - -.02, CI95% - .00 - -.06). Smaller head circumference was associated with more consecutive days with temperatures >30°C at preconception (*B* -.01, CI95% -.00 - .03) and with MEI (*B* -.01, CI95% -.01 - .05) in the first trimester. Conclusions: Overall, exposure to adverse weather during pregnancy is associated with more negative birth outcomes. Therefore, climate change adaptation efforts should pay greater attention to limiting pregnant women’s exposure to adverse weather.

**Key messages:** *What is already known on this topic:*

- The effect of adverse weather on the incidence of some pregnancy disorders has been well-documented, but less is known about the effects on health at birth of intrauterine exposure to adverse climate conditions.
- Health at birth is an important determinant of physical development, cognitive achievement, and work outcomes.
- Global warming has the potential to negatively affect millions with great impacts expected on public health.

*What this study adds:*

- This study highlights that adverse weather conditions are associated with more negative birth outcomes, which may have lasting unfavorable health impact.
- In a large registry-based study, we found that more consecutive days with higher temperature, poor harvest and an interaction between insufficient rain and poor harvest contributed to lower birthweight.
- More days with elevated temperature was associated with shorter neonatal length, whereas more consecutive days with elevated temperature was linked to smaller head circumference.
- Apgar scores were hardly affected by adverse weather conditions.
- Particularly for Tanzania, our results suggest long-term climate-driven health and economic challenges.

*How this study might affect research, practice or policy:*

- The study findings may help policy makers to prioritize and develop programs aiming to reduce climate stress whilst increasing medical preparedness and care for adverse birth outcomes.
- Mitigation of climate effects on pregnant women should receive greater attention than has hitherto been the case in climate change adaptation policy.
- The study points to the need to examine the precise biological mechanisms linking pregnancy and birth outcomes to different weather conditions.

## 1. INTRODUCTION

There is strong agreement in the scientific community that climate change is causing more frequent extreme weather events including high temperatures, droughts, and excessive rainfall.(1) Such events have particularly severe implications for low-income countries due to their low adaptation capacity and higher vulnerability, given their increased exposure, to climate change.(2-4) One important implication concerns public health.(5, 6) Various studies have in fact found heat and cold waves to be linked to higher mortality (7, 8) and spontaneous fetal death rates.(9) However, poor infant health at birth, another possible climate-driven effect, has not yet been investigated in depth(10). It is important to fill this knowledge gap, especially because adverse birth outcomes can have long-lasting consequences to public health systems.

Previous research shows that maternal stress during pregnancy is associated with more complications and suboptimal infant health.(11, 12) This can be explained by the fact that the intrauterine environment can be influenced by external factors(11) that can contribute to lasting fetal structural, functional and metabolic alterations, possibly leading to overt disease later in life.(13) External factors potentially increasing the risk of pregnancy complications include climate change and associated extreme local weather events.(14)

Besides stress, in the sense of biological stressors and self-reported distress, at least two other mechanisms, potentially associated with climate change, might impact offspring health at birth. First, adverse climatic conditions may increase the incidence of infections, since such conditions influence the survival and reproduction of vectors, pathogens and hosts.(15) Second, they can negatively affect access to and consumption of food.(16) Poor birth outcomes, in turn, may negatively affect future health, educational attainment and economic trajectories.(17)

In an ideal world, the empirical assessment of weather effects on birth outcomes should be based on postpartum data for the millions of newborns from low-income countries, where pregnant women are subject to strong variation in exposure to climatic conditions and lack the ability to mitigate their vulnerability in this regard. Yet, precisely in these countries, systematic individual-level data on postpartum infant health over a longer period of time are extremely scarce. For the present study, we were able to obtain access to unique registry data from the Kilimanjaro Christian Medical Centre (KCMC, N=30,068 deliveries), a large hospital in Tanzania.

Few studies reporting data from different sources have explored predictors of birth outcomes in Tanzania or other low-income countries, and the existing ones focus mainly on maternal HIV status,(18, 19) vitamin deficiency,(20) malaria,(21) and pregnancy-related characteristics(22-24). None have examined whether and how environmental conditions are associated with maternal stress and gestational health. Tanzania is a highly interesting country in this regard: It experiences strong weather variability and half of its population lives in poverty(25), with limited means to shield against climate stressors. In the present study, our aims are twofold: first, to examine the extent to which weather conditions namely rainfall, temperature, multivariate ENSO (El Niño Southern Oscillation) index (MEI), effective drought index (EDI), and harvest are associated with neonatal birthweight, length, head circumference, and Apgar scores. Second, we aim to evaluate changes in weather conditions across the studied period. We examine this relationship based on 15 years of data, with weather conditions measured in each pregnancy trimester and a preconception trimester.

## 2. METHODS

### Sample

Our analysis relies on maternal data collected from the KCMC medical birth registry from 2001-2015. All participants providing data on the aforementioned period and fulfilling the inclusion criteria were included. KCMC is is a University teaching hospital with an average of 4,000 births each year (26). The sample includes women delivering singleton babies at the KCMC, which - together with four other zone referral hospitals - serves a population of approximately 15 million in northern Tanzania.

### Medical Birth Registry

A standardized questionnaire-interview was completed for every delivery by trained nurse-midwives who collected data within 24 hours after birth. Interview data included parents’ sociodemographic characteristics, maternal health before and during present pregnancy, delivery characteristics, and maternal reproductive history. Furthermore, women were asked about tobacco, alcohol and medication use. Newborns’ characteristics were extracted from medical records including sex, date and time of delivery, gestational age, abnormal conditions (e.g. birth defects), and Apgar scores. From the medical records, we extracted information regarding birth outcomes: birthweight, measured in kilograms, length, measured in centimeters, head circumference, measured in centimeters and Apgar scores at 1, 5 and 10 minutes, recorded on a scale from 1-10.

We had access to the complete dataset with participants who delivered at KCMC from 2001-2015. However, we focus on examining lower-risk pregnant women delivering at term. By excluding high-risk pregnancies, we avoid the possibility of family-wise type 1 error that may emanate from including a more ill-affected population and our results are expected to be a more valid representation of the effects of interest. Criterion used to exclude higher-risk pregnant women was major obstetric complications (e.g. preeclampsia, bleeding, and infections) in the current pregnancy as defined by the midwives’ reports. We also excluded records with premature birth (< 36 gestational weeks), multiple births, neonatal death and emergency caesarean section, to prevent overestimation of high-risk pregnancies. Details on the sampling strategy can be found in Suppl. Figure 1.

### Weather Data

The analysis of the weather data focuses on six regions in northern Tanzania (from northwest to southeast: Arusha, Kilimanjaro, Manyara, Moshi, Singida and Dodoma; see Suppl.Fig 2). The results presented in this study represent the average for all six regions. The climate in Northern Tanzania is arid with an annual bi-modal rainfall distribution with two distinct rainy seasons (long and short rains), which gradually transform into an unimodal rainfall distribution predominant in the geographical centre of Tanzania (see Suppl. Fig 3 and 4). An overview of all environmental variables used in this study can be seen in Table 1.

**Table 1.**
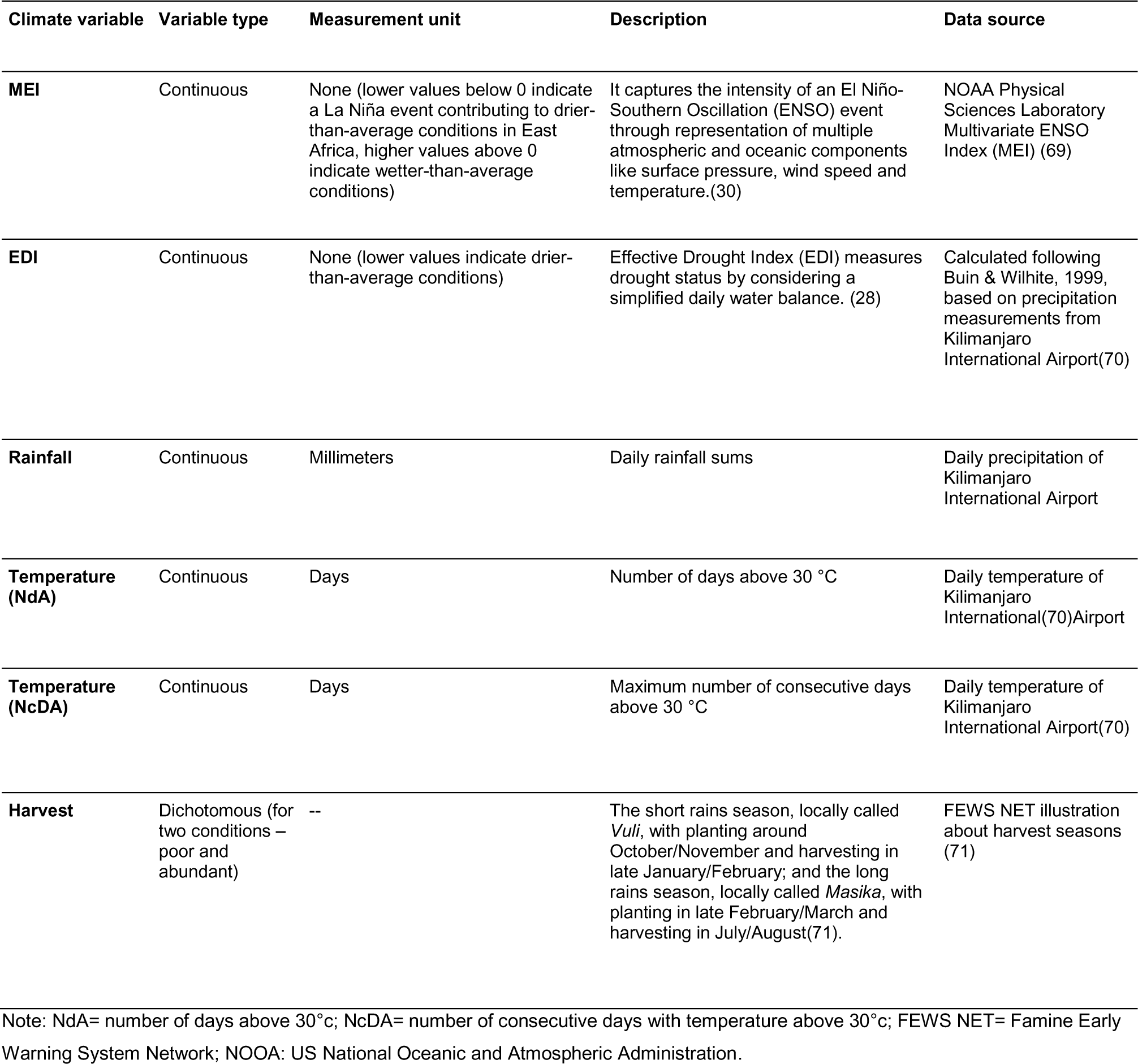
Weather conditions included as predictors in regression analyses.

Transition of seasons is captured by the weather station - Kilimanjaro International Airport (KIA) - data (Daily Surface Summary data [56]) and by the re-analysis data (daily rainfall estimates(27)). As the exact location of pregnant women within their reported region is unknown, the precipitation signal of KIA was used to calculate drought statistics for the complete area, including temperatures, in order to not lose rainfall variability due to averaging over each region. The daily rainfall sums measured at KIA [56] were used to calculate the Effective Drought Index (EDI) following Byun and Wilheit (28). In this study, we collect indicators of heat stress (e.g. excessive heat exposure), humid conditions (which favour flooding and the propagation of pathogens in water) and dry conditions (which correlate with poor harvests and poor nutritional food for the rural population), which in combination provide a good assessment of weather adversity. Seasonality was removed from all weather indicators. Generally, single-parameter indicators for heat (e.g. temperature) are less suitable for assessing temperature stress(29) in humans, but due to its simplicity and availability are often applied in climate science literature (30, 31).Therefore, we also adopted other heat indicators such as the Multivariate ENSO Index (MEI) and EDI.

MEI is an indicator for a large-scale weather pattern, namely the El Niño/Southern Oscillation (ENSO) conditions(30). Prolonged periods with positive MEI are called El Niño events and are associated with wetter than average conditions in east Africa, whereas periods with negative MEI, called La Niña events, are associated with dryer than average conditions. The regional impact of ENSO on the climate and on agricultural production in East Africa is well known and its use is recommended as an early warning system for socio-economic disasters. EDI is an indicator for hydrological drought derived from daily precipitation data. Drought has been found to be a driver for malnutrition with corresponding impacts on health (32) and mental health (33).The health impacts caused by drought are more severe for low-income communities with low resilience (34). Given our interest in high heat periods, temperature was measured as the number of days above 30°C (NdA) and the number of consecutive days above 30°C (NcDA) per trimester.Higher than average and extreme rainfall events are associated with loss of life through flooding, the potential spread of infectious diseases through water (35, 36) and reduced water quality(37). Contrarily, lower than average rainfall can negatively impact plant health, especially in Northern Tanzania(38). Since we were interested in understanding weather effects in all gestational phases, weather conditions were calculated for the preconception trimester and the first, second, and third trimesters of each pregnancy by using as reference the delivery dates. Season-dependent data (e.g., harvest) were used based on the local season (e.g. *Masika* and *Vuli*) and not on gestational trimesters. Table 1 describes in detail the climate variables included in our analysis. In order to facilitate interpretation, the variables rainfall, EDI and MEI were Z-transformed prior to analysis.

### Covariates

Covariates were selected based on relevant literature and on significant correlations with the outcome variables. We included maternal age (continuous, measured in years), maternal education (binary, higher education vs. others), maternal occupation (binary, professional vs. others), marital status (binary, married vs. others), maternal body weight (continuous, measured in kilograms), maternal body height (continuous, measured in centimeters), mother received prenatal care (binary, yes vs. no), maternal alcohol consumption in pregnancy (binary, yes vs. no), maternal diseases before pregnancy (binary, yes vs. no), maternal diseases during pregnancy (binary, yes vs. no), food reserves (continuous, measured in tons). We did not include medication use, smoking, water access and current use of HIV treatment as covariates due to their lack of variability within the sample (all > 92%).

### Statistical Analyses

Statistical analyses were performed using the IBM Statistical Package for the Social Sciences (SPSS Version 24 for Windows) and in R (version 4.2.2). Since most variables were skewed, we applied log transformation using the formula lg10 (value +1) (39). Visual inspections and Kolmogorov–Smirnov tests showed that variables were still not normally distributed, even though the skewness of the data was improved. Therefore, we further used non-parametric tests with non-transformed values (e.g. Spearman correlation). In addition, the logarithm transformed dependent variables were used in the regression models in which the unstandardized residuals were checked for normality. We analyzed whether weather variables were correlated with birth outcomes, as captured by Spearman correlations. We then examined the extent to which each weather condition was associated with each birth outcome after adjusting for covariates. Interaction terms between sufficient rainfall and fruitful harvest as well as insufficient rainfall and poor harvest were estimated and also considered as predictors. We combined these interdependent variables to examine whether they would exert a stronger effect on birth outcomes compared to its single variables. For this purpose, we used linear multivariate regression analysis. While modelling, diagnostics were undertaken to improve model specification, including testing for multicollinearity, but none was found (VIF < 3). Then, we estimated the implications of changes in weather conditions across the studied period based on Friedman tests and Wilcoxon signed-rank post hoc tests. Bonferroni correction was used in all tests (see tables footnotes).

The ethics protocol for this study was approved by The National Institute for Medical Research (NIMR/HQ/R.8a/Vol. IX/2587) and by the KCM College Research Ethics. The Ethics Committee of the Faculty of Arts, University of Zurich, also granted ethical approval (Nr.17.8.8). A data transfer agreement between the KCMC and the University of Zurich was signed in November 2017. The study protocol followed ethical guidelines and has been performed in accordance with the Declaration of Helsinki. Prior to the interview, women received information about the data registry aims and assurance of data privacy. All participants provided oral informed consent before being interviewed and could choose not to participate or not reply to certain questions.

## 3. RESULTS

Sample characteristics can be seen in Table 2. Slightly more participants (51.4%) gave birth to male babies, with a median birthweight of 3185 grams (IQR:600), a mean length of 49 cm (IQR:71), and a mean head circumference of 34 cm (IQR:24). We identified low birthweight (< 2500 grams) in 6.6% of babies, with 10.4% and 8.3% displaying, respectively, shorter-than-average length and head circumference. More than half of participants stated that they had suffered from a serious illness before or during pregnancy (e.g. malaria, cardiac problems).

**Table 2.** Sociodemographic characteristics of the sample.

| Variable | Median (IQR) | % |
| --- | --- | --- |
| Maternal age | 27(9) |  |
| Maternal education |  |  |
| None |  | 1.0 |
| Primary (years 1-7) |  | 52.9 |
| Secondary (years 8-11) |  | 10.6 |
| Higher (12+) |  | 35.5 |
| Marital status |  |  |
| Married |  | 87.9 |
| Single |  | 11.4 |
| Other |  | 0.7 |
| Maternal weight (kg) | 60 (16) |  |
| Maternal height (cm) | 160 (9) |  |
| Received antenatal care (Y) |  | 99.7 |
| *Smoking in pregnancy (N) |  | 92.4 |
| Alcohol in pregnancy (N) |  | 70.3 |
| *Medication in pregnancy (Y) |  | 95.5 |
| Diseases before pregnancy (Y) |  | 62.5 |
| Diseases in pregnancy (Y) |  | 51.3 |
| Baby's sex (M) |  | 51.4 |
| Baby's weight (kg) | 3185.0 (600) |  |
| Baby's length (cm) | 49 (71) |  |
| Baby's head circumference (cm) | 34 (24) |  |
| *Distance to water < 1 km |  | 98.1 |
| Apgar score at 1 min | 9 (10) |  |
| Apgar score at 5 min | 10 (10) |  |
| Apgar score at 10 min | 10 (10) |  |

Note: IQR: Interquartile range, kg= kilograms, cm= centimeters, Y= yes, N= no, M=male; \*Variables not controlled for in analyses due to low variability within the sample

Changes in the development of birth outcomes and birth rates were also seen over time (see Supplementary Figure 5). Apgar scores increased between measurements, whereas other birth outcomes remained relatively stable over the study period. In our sample, a stable birth rate (Supplementary Figure 6) is seen over time, with a slight decrease in the birth rate in 2004 and a greater decline between 2011 and 2012, a trend observed in the whole country (40).

### Correlations between weather variables, covariates and birth outcomes

Spearman correlations revealed that most birth outcomes were significantly correlated with maternal sociodemographic and medical information (range *r_s_* -.028 - .255, *p*≤. 01), and food reserves (range *r_s_* - .019 - .030, p≤. 01). Neonatal birthweight, length and head circumference were correlated with each other (range *r_s_* .48 - .53, *p*≤. 01) as well as with Apgar scores (range *r_s_* .08 - .10, *p*≤. 01). Correlations between weather conditions and birth outcomes can be seen in Suppl.Table 1.

### Associations between weather conditions, covariates and birth outcomes

Data for weather conditions had approximately normally distributed residuals. Findings suggest a relationship between higher maternal education and increased neonatal birthweight (B .01, CI95% .00 .03, p*≤*.002) as well as a positive association between stored food reserves and neonatal birthweight (see Table 3). Higher neonatal birthweight was associated with increased EDI and rainfall, whereas lower birthweight was related to higher MEI, higher NcDA (number of consecutive days above 30°C), poor harvest and the interaction between insufficient rain and poor harvest. The coefficients showed that a one standard deviation increase in the interaction term reduced birthweight by 0.2 kilograms. The interaction term including abundant harvest and sufficient rain revealed null findings.

**Table 3.** Linear multivariate regression models examining effects of weather conditions and covariates on neonatal birthweight.

|  | Rainfall |  | Temperature NdA 30°C |  | Temperature NcDA 30°C |  | MEI |  | Poor harvest x insufficient rain |  | EDI |  |
| --- | --- | --- | --- | --- | --- | --- | --- | --- | --- | --- | --- | --- |
|  | B | CI95% | B | CI95% | B | CI95% | B | CI95% | B | CI95% | B | CI95% |
| *Independent Variables* |  |  |  |  |  |  |  |  |  |  |  |  |
| Maternal age | .00 | .00-.00 | .00 | .00-.00 | .00 | .00-.01 | .00 | .00-.01 | .00 | .00-.00 | .00 | .00-.01 |
| Maternal education | .01* | .00-.02 | .01* | .00-.02 | .01* | .00-.05 | .01 | .00-.02 | .01* | .00-.02 | .01* | .00-.02 |
| Maternal weight | .01* | .01-.04 | .01* | .01-.05 | .01* | .01-.05 | .01* | .00-.05 | .01* | .01-.05 | .01* | .01-.03 |
| Maternal height | .01* | .01-.04 | .01* | .01-.03 | .01* | .01-.03 | .01* | .00-.03 | .01* | .01-.03 | .01* | .01-.05 |
| Antenatal care | .12 | .00-.24 | .11 | .00-.23 | .11 | .00-.23 | .12 | .00-.24 | .11 | -.01-.23 | .12 | .00-.24 |
| Diseases before pregnancy | .01 | .00-.02 | .01 | -.01-.02 | .01 | -.01-.02 | .01 | -.01-.02 | .00 | -.01-.02 | .01 | -.01-.02 |
| Diseases in pregnancy | -.05* | -.04-.05 | -.05* | -.03-.01 | -.05* | -.04-.01 | -.05* | -.03-.02 | -.05* | -.03-.02 | -.05* | -.05-.03 |
| Alcohol in pregnancy | .02* | .00-.05 | .03* | .01-.05 | .02* | .01-.04 | .02* | .01-.04 | .02* | .01-.04 | .02* | .01-.04 |
| Baby's length | .13* | .10-.15 | .13* | .10-.14 | .13* | .10-.14 | .13* | .10-.14 | .13* | .10-.14 | .13* | .10-.14 |
| Baby's head circumference | .22* | .20-.25 | .21* | .20-.24 | .21* | .20-.22 | .22* | .20-.22 | .21* | .20-.25 | .22* | .20-.22 |
| Food reserves | .01* | .00-.05 | .02* | .00-.05 | .01* | .00-.04 | .01* | .00-.05 | .01* | .00-.05 | .01* | .00-.04 |
| Rainfall |  |  |  |  |  |  |  |  |  |  |  |  |
| Preconception | .00 | -.01-.01 |  |  |  |  |  |  |  |  |  |  |
| 1<sup>st</sup> trimester | .01 | .00-.01 |  |  |  |  |  |  |  |  |  |  |
| 2<sup>nd</sup> trimester | .01* | .00-.04 |  |  |  |  |  |  |  |  |  |  |
| 3<sup>rd</sup> trimester | .01 | .00-.04 |  |  |  |  |  |  |  |  |  |  |
| Temperature NdA 30°C |  |  |  |  |  |  |  |  |  |  |  |  |
| Preconception |  |  | .00 | .00-.00 |  |  |  |  |  |  |  |  |
| 1<sup>st</sup> trimester |  |  | .00 | .00-.00 |  |  |  |  |  |  |  |  |
| 2<sup>nd</sup> trimester |  |  | .00 | .00-.00 |  |  |  |  |  |  |  |  |
| 3<sup>rd</sup> trimester |  |  | .00 | .00-.00 |  |  |  |  |  |  |  |  |
| Temperature NcDA 30°C |  |  |  |  |  |  |  |  |  |  |  |  |
| Preconception |  |  |  |  | .00 | .00-.05 |  |  |  |  |  |  |
| 1<sup>st</sup> trimester |  |  |  |  | -.00 | -.00-.01 |  |  |  |  |  |  |
| 2<sup>nd</sup> trimester |  |  |  |  | -.00 | -.00-.01 |  |  |  |  |  |  |
| 3<sup>rd</sup> trimester |  |  |  |  | -.01* | -.00-.05 |  |  |  |  |  |  |
| MEI |  |  |  |  |  |  |  |  |  |  |  |  |
| Preconception |  |  |  |  |  |  | -.03* | -.01-.02 |  |  |  |  |
| 1<sup>st</sup> trimester |  |  |  |  |  |  | .03 | .01-.04 |  |  |  |  |
| 2<sup>nd</sup> trimester |  |  |  |  |  |  | -.01 | -.00-.01 |  |  |  |  |
| 3<sup>rd</sup> trimester |  |  |  |  |  |  | .00 | -.01-.01 |  |  |  |  |
| Poor harvest x Insufficient rain |  |  |  |  |  |  |  |  |  |  |  |  |
| Insufficient rain x poor harvest |  |  |  |  |  |  |  |  | .20* | .13-.26 |  |  |
| Poor harvest |  |  |  |  |  |  |  |  | -.13* | -.10-.08 |  |  |
| EDI |  |  |  |  |  |  |  |  |  |  |  |  |
| Preconception |  |  |  |  |  |  |  |  |  |  | .00 | .00-.01 |
| 1<sup>st</sup> trimester |  |  |  |  |  |  |  |  |  |  | .01 | .00-.02 |
| 2<sup>nd</sup> trimester |  |  |  |  |  |  |  |  |  |  | .01* | .00-.03 |
| 3<sup>rd</sup> trimester |  |  |  |  |  |  |  |  |  |  | .01 | .00-.02 |

Note: Each of the weather conditions was separately modelled with linear regressions. The table displays results from all models. Significance (after Bonferroni correction) is indicated with \* for $p \leq .002$ . NdA 30 °C= Number of days above 30°C; NcDA = Number of consecutive days above 30°C, EDI= Effective drought index; MEI= Multivariate ENSO (El Niño Southern Oscillation) index. CI95%= Confidence Interval.

Shorter neonatal length is associated with higher NdA (number of days above 30°C) and the interaction between insufficient rain and poor harvest, whereas longer length is associated with higher EDI and increased rainfall (see Table 4). The coefficients show that a one standard deviation increase in NdA, for instance, is associated with a decrease in neonatal length by 0.01-0.02 cm, while a one standard deviation increase in the interaction term is associated with a decrease in length by 0.05 cm. The interaction term including abundant harvest and sufficient rain showed null-findings.

**Table 4.** Linear multivariate regression models examining effects of weather conditions and covariates on neonatal length.

|  | Rainfall |  | Temperature NdA 30°C |  | Temperature NcDA 30°C |  | MEI |  | Poor harvest x insufficient rain |  | EDI |  |
| --- | --- | --- | --- | --- | --- | --- | --- | --- | --- | --- | --- | --- |
|  | B | CI95% | B | CI95% | B | CI95% | B | CI95% | B | CI95% | B | CI95% |
| *Independent Variables* |  |  |  |  |  |  |  |  |  |  |  |  |
| Maternal age | -.00 | -.00-.01 | -.00 | -.00-.01 | -.00 | -.00-.01 | -.00 | -.00-.01 | -.00 | -.00-.01 | -.00 | -.00-.01 |
| Maternal education | .00 | .00-.02 | -.00 | -.00-.01 | -.00 | -.00-.01 | .00 | .00-.03 | -.00 | .00-.01 | .00 | .00-.03 |
| Maternal weight | -.00 | -.00-.01 | -.00 | -.00-.01 | -.00 | -.00-.02 | -.00 | -.00-.01 | -.00 | .00-.01 | -.00 | -.00-.02 |
| Maternal height | .01* | .00-.04 | .01* | .00-.04 | .01* | .00-.04 | .01* | .00-.04 | .01* | .00-.04 | .01* | .00-.05 |
| Antenatal care | -.04 | -.03-.01 | -.04 | -.07-.01 | -.04 | -.03-.01 | -.04 | -.03-.01 | -.04 | -.03-.01 | -.04 | -.03-.01 |
| Diseases before pregnancy | .00 | -.01-.01 | .00 | .00-.01 | .00 | -.01-.00 | .00 | .00-.01 | -.00 | -.00-.00 | .00 | -.01-.01 |
| Diseases in pregnancy | .00 | .00-.01 | .00 | -.01-.02 | -.00 | -.00-.01 | .00 | -.01-.01 | .00 | .00-.01 | .00 | .00-.01 |
| Alcohol in pregnancy | -.00 | -.00-.01 | -.00 | -.00-.01 | -.00 | -.00-.01 | -.00 | -.00-.01 | -.00 | -.00-.01 | -.00 | -.00-.01 |
| Baby's weight | .01* | .00-.04 | .01* | .00-.03 | .02* | .00-.04 | .01* | .00-.04 | .01* | .00-.04 | .01* | .00-.03 |
| Baby's head circumference | .03* | .01-.04 | .03* | .01-.03 | .03* | .03-.05 | .03* | .00-.03 | .03* | .00-.03 | .03* | .00-.03 |
| Food reserves | .00 | .00-.01 | .01* | .00-.03 | .00 | .00-.01 | .00 | .00-.01 | .00 | .00-.01 | .00 | .00-.01 |
| Rainfall |  |  |  |  |  |  |  |  |  |  |  |  |
| Preconception | .00 | .00-.01 |  |  |  |  |  |  |  |  |  |  |
| 1<sup>st</sup> trimester | .01* | .00-.05 |  |  |  |  |  |  |  |  |  |  |
| 2<sup>nd</sup> trimester | -.02 | -.00-.06 |  |  |  |  |  |  |  |  |  |  |
| 3<sup>rd</sup> trimester | -.02 | -.00-.05 |  |  |  |  |  |  |  |  |  |  |
| Temperature NdA 30°C |  |  |  |  |  |  |  |  |  |  |  |  |
| Preconception |  |  | -.01* | -.00-.04 |  |  |  |  |  |  |  |  |
| 1<sup>st</sup> trimester |  |  | -.01* | -.00-.04 |  |  |  |  |  |  |  |  |
| 2<sup>nd</sup> trimester |  |  | -.02* | -.00-.06 |  |  |  |  |  |  |  |  |
| 3<sup>rd</sup> trimester |  |  | -.02* | -.00-.05 |  |  |  |  |  |  |  |  |
| Temperature NcDA 30°C |  |  |  |  |  |  |  |  |  |  |  |  |
| Preconception |  |  |  |  | .00 | .00-.01 |  |  |  |  |  |  |
| 1<sup>st</sup> trimester |  |  |  |  | .00 | .00-.01 |  |  |  |  |  |  |
| 2<sup>nd</sup> trimester |  |  |  |  | .00 | .00-.02 |  |  |  |  |  |  |
| 3<sup>rd</sup> trimester |  |  |  |  | .00 | .00-.01 |  |  |  |  |  |  |
| MEI |  |  |  |  |  |  |  |  |  |  |  |  |
| Preconception |  |  |  |  |  |  | -.00 | -.00-.01 |  |  |  |  |
| 1<sup>st</sup> trimester |  |  |  |  |  |  | .00 | .00-.03 |  |  |  |  |
| 2<sup>nd</sup> trimester |  |  |  |  |  |  | -.01 | -.01-.00 |  |  |  |  |
| 3<sup>rd</sup> trimester |  |  |  |  |  |  | .00 | .00-.05 |  |  |  |  |
| Insufficient rain x poor harvest |  |  |  |  |  |  |  |  |  |  |  |  |
| Insufficient rain x poor harvest |  |  |  |  |  |  |  |  | -.05* | -.02-.03 |  |  |
| Poor harvest |  |  |  |  |  |  |  |  | .05* | .03-.06 |  |  |
| EDI |  |  |  |  |  |  |  |  |  |  |  |  |
| Preconception |  |  |  |  |  |  |  |  |  |  | .01* | -.01-.04 |
| 1<sup>st</sup> trimester |  |  |  |  |  |  |  |  |  |  | .01* | -.01-.03 |
| 2<sup>nd</sup> trimester |  |  |  |  |  |  |  |  |  |  | .02 | .00-.05 |
| 3<sup>rd</sup> trimester |  |  |  |  |  |  |  |  |  |  | .13 | .00-.10 |

Note: Each of the weather conditions was separately modelled with linear regressions. The table displays results from all models. Significance (after Bonferroni correction) is indicated with \* for $p \leq .002$ . NdA 30 °C = Number of days above 30°C; NcDA = Number of consecutive days above 30°C, EDI= Effective drought index; MEI= Multivariate ENSO (El Niño Southern Oscillation) index. CI95% = Confidence interval.

We also found that neonatal smaller head circumference is significantly associated with NcDA and higher MEI, whereas larger head circumference is associated with EDI and the interaction between insufficient rain and poor harvest (see Table 5). One standard deviation increase in MEI was associated with a decrease in head circumference by 0.01 cm. We found no significant effects of the interaction between abundant harvest and sufficient rain on head circumference.

**Table 5.** Linear multivariate regression models examining effects of weather conditions and covariates on neonatal head circumference.

|  | Temperature NDA<br>30°C |  | Temperature NcDA<br>30°C |  | MEI |  | Poor harvest x<br>insufficient rain |  | EDI |  |
| --- | --- | --- | --- | --- | --- | --- | --- | --- | --- | --- |
|  | B | CI95% | B | CI95% | B | CI95% | B | CI95% | B | CI95% |
| *Independent Variables* |  |  |  |  |  |  |  |  |  |  |
| Maternal age | -.00 | -.00-.01 | -.01 | .01-.03 | -.001 | -.00-.01 | -.001 | -.00-.02 | -.00 | -.00-.01 |
| Maternal education | -.01 | -.00-.00 | -.00 | -.00-.01 | -.000 | -.00-.01 | -.001 | -.00-.01 | -.01 | -.00-.01 |
| Maternal occupation | -.00 | -.00-.00 | -.01 | -.00-.01 | -.000 | -.00-.01 | -.001 | -.00-.02 | -.00 | -.00-.01 |
| Marital status | .00 | .00-.01 | .00 | .00-.01 | .002 | .00-.07 | .000 | .00-.03 | .00 | .00-.05 |
| Maternal weight | -.00 | -.00-.03 | -.00 | -.00-.02 | -.000 | -.00-.03 | -.001 | -.00-.04 | -.00 | -.00-.03 |
| Maternal height | -.02* | -.00-.01 | -.01* | -.00-.00 | -.001* | -.00-.05 | -.002* | -.00-.03 | -.02* | -.00-.01 |
| Antenatal care | .04 | .01-.07 | .04 | .01-.07 | .004 | .01-.06 | .004 | .01-.07 | .04 | .01-.07 |
| Diseases before pregnancy | .01 | .00-.05 | -.00 | -.00-.01 | .001 | .00-.03 | .001 | .00-.04 | -.00 | -.00-.02 |
| Diseases in pregnancy | .01* | .00-.01 | .01 | .00-.01 | .000 | .00-.01 | .001* | .00-.03 | .01* | .00-.05 |
| Alcohol in pregnancy | -.03 | .00-.06 | -.01 | -.00-.01 | -.001 | -.00-.03 | -.000 | -.00-.02 | -.00 | -.00-.01 |
| Baby's weight | .01* | .00-.03 | .01* | .00-.03 | .002* | .00-.04 | .001* | .00-.03 | .02* | .00-.05 |
| Baby's length | .02* | .02-.08 | .02* | .02-.07 | .002* | .02-.05 | .002* | .01-.04 | .01* | .01-.04 |
| Food reserves | -.01* | -.00-.06 | .00* | .00-.00 | -.001* | -.00-.02 | -.001* | -.00-.03 | -.00* | -.00-.03 |
| Temperature Nda 30°C |  |  |  |  |  |  |  |  |  |  |
| Preconception | -.01 | -.00-.00 |  |  |  |  |  |  |  |  |
| 1<sup>st</sup> trimester | .01 | .00-.00 |  |  |  |  |  |  |  |  |
| 2<sup>nd</sup> trimester | -.01 | -.00-.00 |  |  |  |  |  |  |  |  |
| 3<sup>rd</sup> trimester | .01 | .00-.00 |  |  |  |  |  |  |  |  |
| Temperature NcDA<br>30°C |  |  |  |  |  |  |  |  |  |  |
| Preconception |  |  | -.01* | -.00-.03 |  |  |  |  |  |  |
| 1<sup>st</sup> trimester |  |  | -.00 | -.00-.01 |  |  |  |  |  |  |
| 2<sup>nd</sup> trimester |  |  | -.00 | -.00-.02 |  |  |  |  |  |  |
| 3<sup>rd</sup> trimester |  |  | .00 | .00-.01 |  |  |  |  |  |  |
| MEI |  |  |  |  |  |  |  |  |  |  |
| Preconception |  |  |  |  | .01 | .00-.04 |  |  |  |  |
| 1<sup>st</sup> trimester |  |  |  |  | -.01* | -.01-.05 |  |  |  |  |
| 2<sup>nd</sup> trimester |  |  |  |  | .00 | .00-.02 |  |  |  |  |
| 3<sup>rd</sup> trimester |  |  |  |  | .00 | .00-.03 |  |  |  |  |
| Poor harvest x Insufficient rain |  |  |  |  |  |  |  |  |  |  |
| Insufficient rain x poor harvest |  |  |  |  |  |  | -.06* | -.05-.02 |  |  |
| Poor harvest |  |  |  |  |  |  | .03* | .01-.04 |  |  |
| EDI |  |  |  |  |  |  |  |  |  |  |
| Preconception |  |  |  |  |  |  |  |  | -.00 | -.00-.02 |
| 1<sup>st</sup> trimester |  |  |  |  |  |  |  |  | .00 | .00-.05 |
| 2<sup>nd</sup> trimester |  |  |  |  |  |  |  |  | .01* | .00-.04 |
| 3<sup>rd</sup> trimester |  |  |  |  |  |  |  |  | .01* | .00-.05 |

Note: Each of the six weather conditions was separately modelled with linear regressions. The table displays results from all models Significance (after Bonferroni correction) is indicated with \* for $p \leq .002$ , Nda 30 °C = Number of days above 30°C; NcDA = Number of consecutive days above 30°C, EDI= Effective drought index; MEI= Multivariate ENSO (El Niño Southern Oscillation) index; min = minutes. CI95% = Confidence interval.

In contrast to the findings discussed above, Apgar scores at 1, 5 and 10 min were only weakly associated with weather conditions. We only found a significant relationship between increased poor harvest and lower Apgar scores only at 1 min (*B* -.05, CI95% -.00 to .01).

### Weather changes over time

To examine the implications of alterations in weather conditions over time, we conducted a Friedman test, and to distinguish effects between gestational trimesters (and the preconception trimester), we additionally conducted a Wilcoxon signed-rank post hoc test. The results of the Friedman test pointed to a significant difference (Bonferroni correction p ≤0.01; four tests) from preconception to the third gestational trimester for the following weather variables: rainfall, χ^2^ (3) 202.07; EDI, χ^2^ (3) 397.09; NdA, χ^2^(3) 65.55; NcDA, χ^2^(3) 75.52; MEI, χ^2^(3) 307.66. In the post hoc analysis (Bonferroni correction p ≤0.01), the following significant differences emerged: rainfall, between preconception and first trimester (*Z* -7.46), second and third trimester (*Z* -5.74), and preconception and third trimester (*Z* -14.55); EDI, between preconception and first trimester (*Z* -8.28), and preconception and third trimester (*Z* -13.56); MEI, between preconception and first trimester (*Z* -6.31) and preconception and third trimester (*Z* - 12.43); NdA, between first and second trimester (*Z* -5.90), second and third trimester (*Z* -3.99), and preconception and third trimester (*Z* -7.13); NcDA, between preconception and first trimester (*Z* -9.82), first and second trimester (*Z* -14.19), second and third trimester (*Z* -13.21) and preconception and third trimester (*Z* -5.92).

### Supplementary analyses

Additional analyses including high-risk pregnancies (e.g. premature birth, multiple births, neonatal death and emergency caesarean section) indicated that the results reported above did not differ when analysing the complete dataset; that is, similar weather effects on birth outcomes were found when jointly analysing high and low-risk pregnancies. A second set of analyses considered potential individual moderation effects of prematurity, neonatal death and emergency c-section on the associations between weather and birth outcomes. Results showed that there was no reliable evidence that these associations were moderated by prematurity, neonatal death or emergency c-section (non-significant results). A third set of analyses considered premature birth, neonatal death and emergency caesarean section as outcomes. Increased NdA at first and third gestational trimester respectively (Exp (B) 1.354, 1.327, p<.001), and poor harvest (Exp (B) 1.267, p<.001) heightened the odds of a neonatal death. Moreover, the regression models predicting c-section showed that increased MEI in the second gestational trimester was 1.08 times more likely to contribute to c-section (p<.001). Finally, high MEI (Exp (B) 1.354 p<.001) in the second gestational trimester and poor harvest (Exp (B) 1.418, p<.001) increased the odds of prematurity. A final set of supplementary analyses controlled for birth year, since a particular event such as a disease outbreak in a specific year could have affected birth outcomes. Results suggest that the associations between weather variables and birth outcomes were significant and similar in magnitude to the complete dataset.

## 4. DISCUSSION

In this study, we aimed to examine the extent to which weather conditions are associated with neonatal outcomes and to evaluate changes in weather conditions across the studied period. Our findings indicate that consecutive days with higher temperature, poor harvest and an interaction between insufficient rain and poor harvest contributed to lower birthweight. More days with elevated temperature was associated with shorter neonatal length, whereas consecutive days with higher temperature was linked to smaller head circumference. Few associations were found between weather conditions and Apgar scores. All weather variables significantly changed between preconception and the three gestational trimesters.

The association of lower birthweight with exposure to higher temperatures during pregnancy is in line with previous literature. (41, 42) In particular, we found that consecutive high temperature days (NcDA) in the third gestational trimester negatively impacted birthweight. This is biologically plausible, as the third trimester is an important time for fetal growth.(43) Since extreme temperatures may affect uterine blood flow and placental exchange necessary for fetal maturation,(44) disrupting this mechanism at this time would have the greatest impact on weight. Conversely, above-average (but not excessive) rainfall may benefit birthweight through cooler temperatures and higher agricultural productivity, improving maternal nutrition and increasing the amount of drinkable water. (45) The association between increased birthweight and rainfall in mid-pregnancy confirms that birthweight sensitivity to weather seems to be concentrated in later pregnancy stages. (46) The combination of two extreme conditions, insufficient rain and poor harvest throughout pregnancy, negatively affected birthweight. Reduced harvests due to natural disasters increase food prices, contributing to food insecurity(47), which can in turn result in undernutrition. (48) However, a secondary pathway through which food insecurity may affect birthweight is maternal stress. (49) For instance, in a previous study, 76% of women from Northern Tanzania identified hunger as one of the most significant stressors in their lives. (50) Maternal stress is a well-established risk factor for adverse birth outcomes.(51) In our sample, maternal higher education was associated with increased birthweight, concurring with previous findings. (52, 53) Less-educated mothers are more likely to have a poor diet due to low income and/or low dietary literacy (53). Additionally, limited education may lead to lower adherence to healthcare messages, resulting in a higher likelihood of delivering a low birthweight baby. (54) Low birthweight has long been associated with adulthood diseases such as hypertension(55), type II diabetes mellitus(56), ischemic cardiomyopathy(57) and respiratory disease(58) among others.

Infants whose mothers had been exposed to excessive heat were shorter in length and had a significantly smaller head circumference. During gestation, women are potentially more affected by heat due to physiological changes altering body temperature regulation. Pregnancy is a hypervolemic state with expanded plasma volume, high cardiac output and decreased vascular resistance (59). Maternal thermoregulation may also be affected by expected weight increases, and by fetal metabolism, which elevates maternal basal metabolic rate (59). Similarly to low birthweight, shorter neonatal length has been strongly related to coronary heart disease later in life, particularly among women (60). Seminal work by Broman(61) showed that head circumference at birth is associated with child intellectual performance, and reflects brain size. Considerable size differences were reported in a study comparing stressed and non-stressed women(62), in which offspring of stressed mothers displayed a 5% smaller brain volume. In particular, heat stress (e.g. excessive heat exposure) in pregnancy has been suggested as a risk factor for teratogenesis. (63) If prenatal stress, including heat stress, diminishes head circumference and thus fetal brain growth, it will have far-reaching public health consequences, since exposure to heat in the Kilimanjaro region is common.

According to our data, weather conditions barely seem to affect Apgar scores. Markers like neonatal head circumference and birthweight were not strongly correlated with Apgar in our study or in others. (64, 65) For instance, Iliodromiti and colleagues found no association between birthweight and Apgar scores in their study encompassing more than 1 million births in Scotland. (64) A potential reason for this could be that Apgar is a method to clinically assess newborn status at birth, focusing on a neonate’s appearance and need for support with breathing(66), whereas birth outcomes represent an infant’s morphology and are more predictive of future health outcomes. (67) Another possible explanation for this lack of association lies in inter-observer variability while attributing Apgar scores. (66) The Apgar describes an infant’s physiological condition at one time point, and certain score components, such as tone or reflex irritability, can be subjective(66). Our findings are relevant in showing that factors such as birth outcomes and weather do not seem to have strong implications for Apgar scores.

Our large sample size with higher statistical power, enabled detection of small significant effects and relationships. However, it is important to note that not all weather conditions here investigated had a significant association with the birth outcomes. For instance, MEI was not significantly associated with neonatal length, rainfall was not related to head circumference, NdA showed no associations with neonatal birthweight, whereas Apgar scores were hardly affected by the environmental variables. Moreover, we adjusted our analyses for multiple testing to correct for occurrence of false positives. This means that our significant findings are not due to the large sample size, but more likely capture the characteristics and variability of the population studied, leading to more reliable results. Although our effect sizes were small, they are representative and generalizable on a population level, increasing our confidence in applying the conclusions to the Northern Tanzania context.

The research presented here has limitations that could serve as a starting point for further inquiry. We did not have information on women’s residential mobility, which might implicate some degree of weather exposure misclassification. However, it has been shown that most women who relocate during pregnancy move within 10 km of their previous residence. (68) Some counterintuitive results, such as higher EDI increasing neonatal birthweight and length, require further study. One approach could be to more systematically consider factors that may moderate the effect of weather conditions on pregnant women and their offspring. For instance, it would be worthwhile to use more detailed data than we had on de facto exposure of women during pregnancy (e.g. time spent indoors/outdoors, use of air conditioning). Similarly, we did not have data on maternal mental health, which is an important risk factor for birth outcomes. Another limitation on which further research should focus concerns the biological mechanisms linking pregnancy and birth outcomes to different weather conditions.

## 5. CONCLUSION

Despite these limitations, which will hopefully be addressed in further research, our findings strongly suggest that climate change is likely to have negative implications for birth outcomes in low-income environments. Mitigation of climate effects on pregnant women should thus receive greater attention than has hitherto been the case in climate change adaptation policy. Our findings may help policy makers to prioritize and develop programs aiming to reduce climate stress whilst increasing medical preparedness and care for adverse birth outcomes.

## Data availability statement

The datasets generated during and/or analysed during the current study are available from the corresponding author on reasonable request.

## Data Availability

All data produced during the current study are available from the corresponding author on reasonable request.

## Acknowledgements

The authors would like to thank Stefan Isler for his assistance with the ethics (IRB) approval procedure and data acquisition; to Josephine Beerli, Hannah Meyerhoff and Berit Barthelmes for research assistance. We acknowledge the KCMC and KCM-College administration for authorizing use of the hospital birth registry and neonatal registry data for this study. We thank the staff of the birth registry department in Norway for ongoing supervision and technical support for the KCMC birth registry. We also thank the birth registry team, obstetrics and gynaecology and paediatric staff at KCMC for their support in data collection. We are also grateful to the participants who delivered at KCMC during the period of data collection and their babies.

## Author Contributions

UE and TB conceived the presented idea. RAC analysed the data. BM processed the environmental data and designed the figures. UE, TS, TB, BM and BM verified the analytical methods and contributed to the interpretation of the results. UE supervised the project. RAC took the lead in writing the manuscript. All authors provided critical feedback and contributed to the final version of the manuscript.

## References

1. Intergovernmental Panel On Climate Change. Climate change 2013: the physical science basis: Working Group I contribution to the Fifth assessment report of the Intergovernmental Panel on Climate Change: Cambridge University Press; 2014.

2. Jones PG, Thornton PK. The potential impacts of climate change on maize production in Africa and Latin America in 2055. Global environmental change. 2003;13(1):51–9.

3. Adenle AA, Ford JD, Morton J, Twomlow S, Alverson K, Cattaneo A, et al. Managing climate change risks in Africa-A global perspective. Ecological Economics. 2017;141:190–201.

4. Niang A, Becker M, Ewert F, Dieng I, Gaiser T, Tanaka A, et al. Variability and determinants of yields in rice production systems of West Africa. Field crops research. 2017;207:1–12.

5. Cheng JJ, Berry P. Health co-benefits and risks of public health adaptation strategies to climate change: a review of current literature. International journal of public health. 2013;58(2):305–11.

6. Garland RM, Matooane M, Engelbrecht FA, Bopape M-JM, Landman WA, Naidoo M, et al. Regional projections of extreme apparent temperature days in Africa and the related potential risk to human health. International journal of environmental research and public health. 2015;12(10):12577–604.

7. Gasparrini A, Guo Y, Hashizume M, Lavigne E, Zanobetti A, Schwartz J, et al. Mortality risk attributable to high and low ambient temperature: a multicountry observational study. Lancet (London, England). 2015;386(9991):369-75.

8. Deschênes O, Greenstone M. Climate change, mortality, and adaptation: Evidence from annual fluctuations in weather in the US. American Economic Journal: Applied Economics. 2011;3(4):152–85.

9. Fukuda M, Fukuda K, Shimizu T, Nobunaga M, Mamsen LS, Yding Andersen C. Climate change is associated with male:female ratios of fetal deaths and newborn infants in Japan. Fertil Steril. 2014;102(5):1364.

10. Molina O, Saldarriaga V. The perils of climate change: In utero exposure to temperature variability and birth outcomes in the Andean region. Economics & Human Biology. 2017;24:111–24.

11. Murray LJ, O’Reilly DP, Betts N, Patterson CC, Davey Smith G, Evans AE. Season and outdoor ambient temperature: effects on birth weight. Obstet Gynecol. 2000;96(5 Pt 1):689-95.

12. Castro RA, Ehlert U, Dainese S, Zimmerman R, La Marca-Ghaemmaghami P. Psychological predictors of gestational outcomes in second trimester pregnant women: associations with daily uplifts. Springer; 2020.

13. Flouris AD, Faught BE, Hay J, Cairney J. Exploring the origins of developmental disorders. Dev Med Child Neurol. 2005;47(7):436.

14. Poursafa P, Keikha M, Kelishadi R. Systematic review on adverse birth outcomes of climate change. J Res Med Sci. 2015;20(4):397–402.

15. Patz JA, Githeko AK, McCarty JP, Hussain S, Confalonieri U, De Wet N. CHAPTER 6: Climate change and infectious diseases. 2003. p. 103-32.

16. Skoufias E, Vinha K. The impacts of climate variability on household welfare in rural Mexico. Popul Environ. 2013;34(3):370–99.

17. Victora CG, Adair L, Fall C, Hallal PC, Martorell R, Richter L, et al. Maternal and child undernutrition: consequences for adult health and human capital. Lancet (London, England). 2008;371(9609):340-57.

18. Coley JL, Msamanga GI, Fawzi MCS, Kaaya S, Hertzmark E, Kapiga S, et al. The association between maternal HIV-1 infection and pregnancy outcomes in Dar es Salaam, Tanzania. BJOG. 2001;108(11):1125–33.

19. Habib NA, Daltveit AK, Bergsjø P, Shao J, Oneko O, Lie RT. Maternal HIV status and pregnancy outcomes in northeastern Tanzania: a registry-based study. BJOG. 2008;115(5):616–24.

20. Fawzi WW, Msamanga GI, Urassa W, Hertzmark E, Petraro P, Willett WC, et al. Vitamins and perinatal outcomes among HIV-negative women in Tanzania. N Engl J Med. 2007;356(14):1423–31.

21. Uddenfeldt Wort U, Warsame M, Brabin BJ. Birth outcomes in adolescent pregnancy in an area with intense malaria transmission in Tanzania. Acta Obstet Gyn Scand. 2006;85(8):949–54.

22. Mosha TCE, Philemon N. Factors influencing pregnancy outcomes in Morogoro Municipality, Tanzania. Tanzan J Health Res. 2010;12(4):249–60.

23. Shirima CP, Kinabo JL. Nutritional status and birth outcomes of adolescent pregnant girls in Morogoro, Coast, and Dar es Salaam regions, Tanzania. Nutrition. 2005;21(1):32–8.

24. Watson-Jones D, Weiss HA, Changalucha JM, Todd J, Gumodoka B, Bulmer J, et al. Adverse birth outcomes in United Republic of Tanzania--impact and prevention of maternal risk factors. Bull World Health Organ. 2007;85(1):9–18.

25. Group WB. Poverty & Equity Brief - Tanzania. 2021.

26. Bergsjo P, Mlay J, Lie RT, Lie-Nielsen E, Shao JF. A medical birth registry at Kilimanjaro Christian Medical Centre. East Afr J Public Health. 2007;4(1):1–4.

27. Portal UFND. RFE (Rainfall Estimate) 2022 [Available from: https://earlywarning.usgs.gov/fews/product/48.

28. Byun H-R, Wilhite DA. Objective quantification of drought severity and duration. Journal of climate. 1999;12(9):2747–56.

29. de Freitas CR, Grigorieva EA. A comparison and appraisal of a comprehensive range of human thermal climate indices. International journal of biometeorology. 2017;61(3):487–512.

30. Wolter K, Timlin MS. El Niño/Southern Oscillation behaviour since 1871 as diagnosed in an extended multivariate ENSO index (MEI. ext). International Journal of Climatology. 2011;31(7):1074–87.

31. Schwingshackl C, Sillmann J, Vicedo-Cabrera AM, Sandstad M, Aunan K. Heat stress indicators in CMIP6: estimating future trends and exceedances of impact-relevant thresholds. Earth’s Future. 2021;9(3):e2020EF001885.

32. Lieber M, Chin-Hong P, Kelly K, Dandu M, Weiser SD. A systematic review and meta-analysis assessing the impact of droughts, flooding, and climate variability on malnutrition. Global Public Health. 2022;17(1):68–82.

33. Edwards B, Gray M, Hunter B. The impact of drought on mental health in rural and regional Australia. Social Indicators Research. 2015;121(1):177–94.

34. Stanke C, Kerac M, Prudhomme C, Medlock J, Murray V. Health effects of drought: a systematic review of the evidence. PLoS currents. 2013;5.

35. Zolotokrylin A, Cherenkova E. Seasonal changes in precipitation extremes in Russia for the last several decades and their impact on vital activities of the human population. Geography, environment, sustainability. 2018;10(4):69–82.

36. Parmenter RR, Yadav EP, Parmenter CA, Ettestad P, Gage KL. Incidence of plague associated with increased winter-spring precipitation in New Mexico. The American journal of tropical medicine and hygiene. 1999;61(5):814–21.

37. Aguilera R, Gershunov A, Benmarhnia T. Atmospheric rivers impact California’s coastal water quality via extreme precipitation. Science of the Total Environment. 2019;671:488–94.

38. Nzabarinda V, Bao A, Xu W, Uwamahoro S, Udahogora M, Umwali ED, et al. A spatial and temporal assessment of vegetation greening and precipitation changes for monitoring vegetation dynamics in climate zones over Africa. ISPRS International Journal of Geo-Information. 2021;10(3):129.

39. Field A. Discovering statistics using SPSS: Sage publications; 2009.

40. United Nations. Wolrd Population Prospects 2019 [Available from: https://population.un.org/wpp/.

41. Chodick G, Flash S, Deoitch Y, Shalev V. Seasonality in birth weight: review of global patterns and potential causes. Hum Biol. 2009;81(4):463–77.

42. Ngo NS, Radley MH. Climate change and fetal health: The impacts of exposure to extreme temperatures in New York City. Environ Res. 2016;144:158–64.

43. Buck Louis GM, Grewal J, Albert PS, Sciscione A, Wing DA, Grobman WA, et al. Racial/ethnic standards for fetal growth: the NICHD Fetal Growth Studies. Am J Obstet Gynecol. 2015;213(4):449.e1-.e41.

44. Browne VA, Julian CG, Toledo-Jaldin L, Cioffi-Ragan D, Vargas E, Moore LG. Uterine artery blood flow, fetal hypoxia and fetal growth. Philos Trans R Soc Lond B Biol Sci. 2015;370(1663):20140068.

45. Rocha R, Soares RR. Water scarcity and birth outcomes in the Brazilian semiarid. J Dev Econ. 2015;112:72–91.

46. Deschenes O, Greenstone M, Guryan J. Climate Change and Birth Weight. Am Econ Rev. 2009;99(2):211–7.

47. Von Braun J. Food and financial crises: Implications for agriculture and the poor: Intl Food Policy Res Inst; 2008.

48. Webb P, Coates J, Frongillo EA, Rogers BL, Swindale A, Bilinsky P. Measuring Household Food Insecurity: Why It’s So Important and Yet So Difficult to Do. J Nutr. 2006;136(5):1404S–8S.

49. Siefert K, Heflin CM, Corcoran ME, Williams DR. Food insufficiency and physical and mental health in a longitudinal survey of welfare recipients. Journal of health and social behavior. 2004;45(2):171–86.

50. Pike IL, Patil CL. Understanding women’s burdens: Preliminary findings on psychosocial health among Datoga and Iraqw women of Northern Tanzania. Cult Med Psychiatry. 2006;30(3):299–330.

51. Dunkel Schetter C, Lobel M. Pregnancy and birth outcomes: A multilevel analysis of prenatal maternal stress and birth weight. In: Baum A, Revenson TA, Singer J, editors. Handbook of health psychology, 2nd ed. New York, NY: Psychology Press; 2012. p. 431-63.

52. Silvestrin S, Silva CHd, Hirakata VN, Goldani AAS, Silveira PP, Goldani MZ. Maternal education level and low birth weight: a meta-analysis. J Pediatr (Rio J). 2013;89(4):339–45.

53. Muula AS, Siziya S, Rudatsikira E. Parity and maternal education are associated with low birth weight in Malawi. Afr Health Sci. 2011;11(1):65–71.

54. Pincus T, Callahan LF. Associations of low formal education level and poor health status: behavioral, in addition to demographic and medical, explanations? J Clin Epidemiol. 1994;47(4):355–61.

55. Wadsworth M, Cripps H, Midwinter R, Colley J. Blood pressure in a national birth cohort at the age of 36 related to social and familial factors, smoking, and body mass. Br Med J (Clin Res Ed). 1985;291(6508):1534-8.

56. Hales CN, Barker DJ, Clark PM, Cox LJ, Fall C, Osmond C, et al. Fetal and infant growth and impaired glucose tolerance at age 64. British medical journal. 1991;303(6809):1019-22.

57. Barker DJ, Osmond C. Infant mortality, childhood nutrition, and ischaemic heart disease in England and Wales. The Lancet. 1986;327(8489):1077-81.

58. Barker DJ, Godfrey K, Fall C, Osmond C, Winter P, Shaheen S. Relation of birth weight and childhood respiratory infection to adult lung function and death from chronic obstructive airways disease. British Medical Journal. 1991;303(6804):671-5.

59. Cunningham FG, Leveno KJ, Bloom SL, Spong CY, Dashe JS, Hoffman BL, et al. Obstetricia de Williams: McGraw Hill Brasil; 2016.

60. Barker DJ, Eriksson JG, Forsén T, Osmond C. Fetal origins of adult disease: strength of effects and biological basis. International journal of epidemiology. 2002;31(6):1235–9.

61. Broman SH, Nichols PL, Kennedy WA. Preschool IQ: Prenatal and early developmental correlates: Routledge; 2017.

62. Lou HC, Hansen D, Nordentoft M, Pryds O, Jensen F, Nim J, et al. Prenatal stressors of human life affect fetal brain development. Dev Med Child Neurol. 1994;36(9):826–32.

63. Soultanakis HN. Aquatic Exercise and Thermoregulation in Pregnancy. Clin Obst Gynecol. 2016;59(3):576–90.

64. Iliodromiti S, Mackay DF, Smith GCS, Pell JP, Nelson SM. Apgar score and the risk of cause-specific infant mortality: a population-based cohort study. Lancet (London, England). 2014;384(9956):1749-55.

65. Costa T, Mota A, Duarte S, Araujo M, Ramos P, Machado H. Predictive Factors of Apgar Scores below 7 in Newborns: Can We Change the Route of Current Events. J Anesth Clin Res. 2016;7(672):2.

66. Committee on Obstetric Practice. ACOGAmerican Academy of PediatricsCommittee on Fetus and Newborn, ACOG. ACOG Committee Opinion. Number 644, Oct 2015 (replaces No. 333, May 2006): The Apgar score. Obstet Gynecol. 2015;126(4):E52-E5.

67. Barker DJ. Adult consequences of fetal growth restriction. Clinical obstetrics and gynecology. 2006;49(2):270–83.

68. Bell ML, Belanger K. Review of research on residential mobility during pregnancy: consequences for assessment of prenatal environmental exposures. J Exp Sci Environ Epidemiol. 2012;22(5):429–38.

69. Laboratory NPS. Multivariate ENSO Index (MEI) 2017 [Available from: https://psl.noaa.gov/enso/mei/.

70. Branch NCS. Global Surface Summary of the Day Data, Version 7 2017 [Available from: https://www.ncei.noaa.gov/maps/daily/

71. MacLeod D. Seasonal predictability of onset and cessation of the east African rains. Weather and climate extremes. 2018;21:27–35.

