## Supplementary Material for "Adverse Weather Conditions can have Negative Effects on Birth Outcomes: Evidence from a birth registry cohort in Tanzania"

**Supplementary Figures Legends**

**Supplementary Figure 1.** Sampling strategy. Note: *These records were excluded to reduce potential effects of typing errors, the most plausible reason for such unrealistic measures.

**Supplementary Figure 2.** Overview of the study region

**Supplementary Figure 3.** Climate in Tanzania

**Supplementary Figure 4.** Monthly temperatures and precipitation

**Supplementary Figure 5.** Development of birth weight, neonatal length and neonatal head circumference from 2001-2015.

**Supplementary Figure 6.** Birth frequency from 2001-2015

**Supplementary Tables Legend**

**Supplementary Table 1.** Spearman correlations between weather conditions and birth outcomes

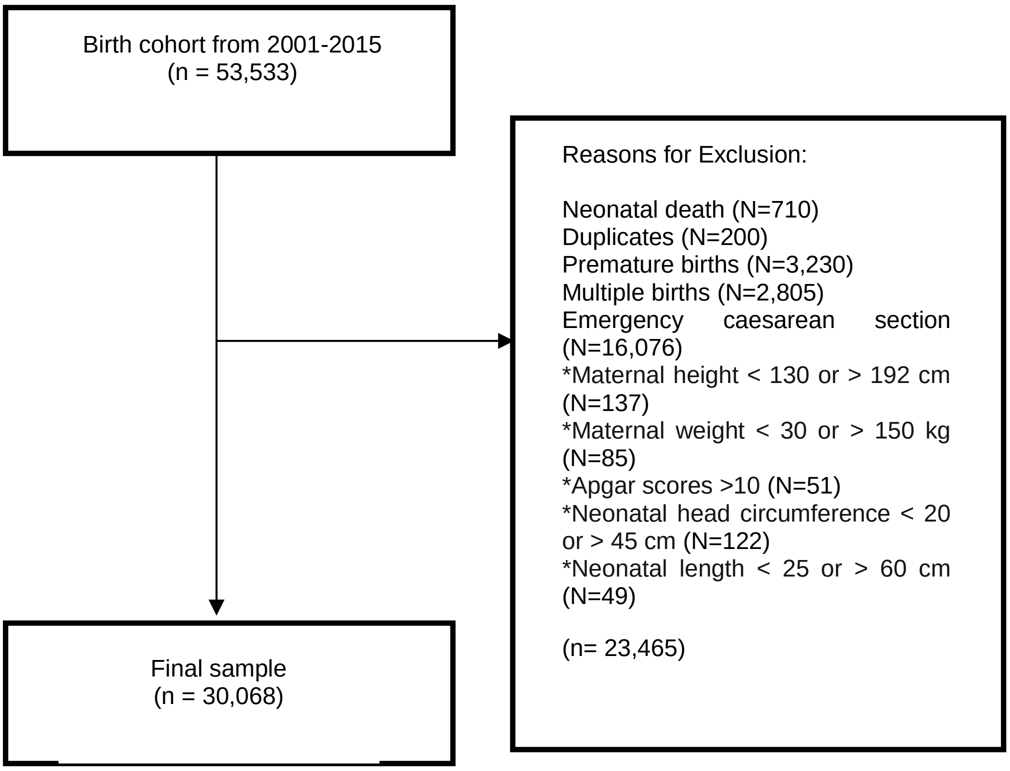


**Supplementary Figure 1**. Sampling Strategy.

Note: *These records were excluded to reduce potential effects of typing errors, the most plausible reason for such unrealistic measures.

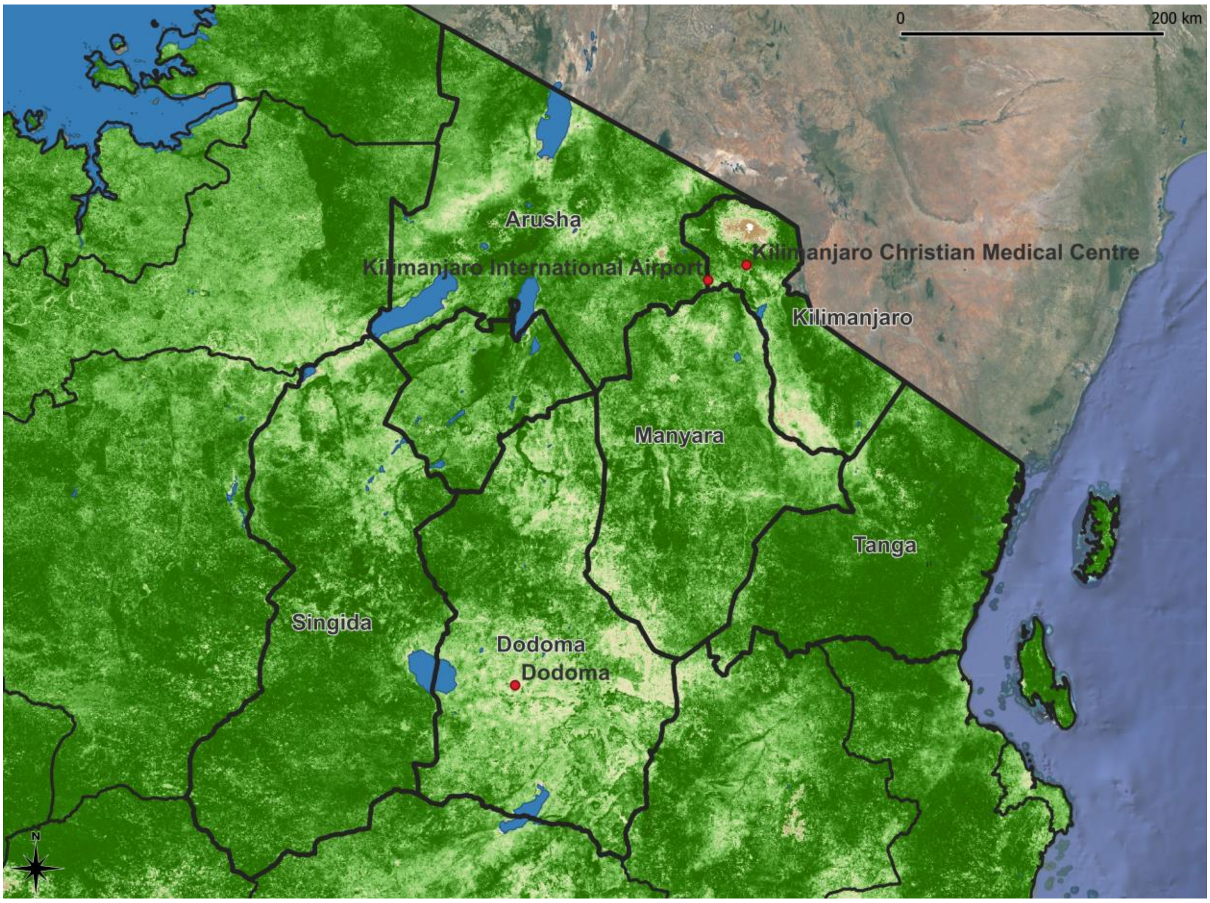


**Supplementary Figure 2.** Overview of the study region

Note: The figure shows the north-east of Tanzania with the location of the KCMC as the northern most red point. In the background an image of January 2001 (rainy season) is shown. Dark green colours indicate high density of green vegetation whereas brown colours indicate sparce or brown vegetation. The nearest weather station from which weather data was available is located approximately 30 km to the south-west of the hospital at Kilimanjaro International Airport.

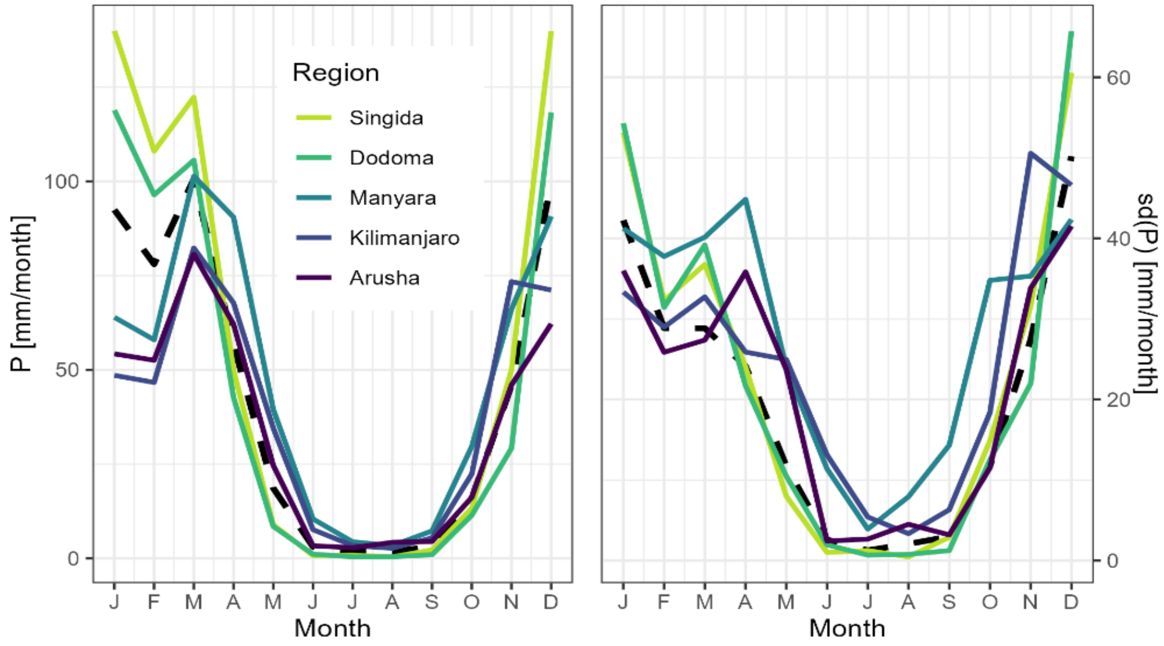


**Supplementary Figure 3.** Climate in Tanzania

Note1: The figure shows the average monthly rainfall pattern for each of the regions in north-east Tanzania that was named as home region by the women giving birth. There is hardly any rainfall between June and September. The two southern regions, Dodoma and Singida, show a unimodal rainfall pattern whereas the northern regions display a clear bi-modal rainfall pattern. Rainfall patterns in the north of the country, where the majority of the pregnancies analysed here occur, are very similar and highly correlated. The living location of the pregnant women is not known accurately. The present study resorts to using station precipitation data from Kilimanjaro International Airport for the calculation of the effective drought index and not the average precipitation sums for each region since due to the size of the regions, the regionally averaged precipitation signal became heavily smoothed and did not represent realistic precipitation patterns. It further allowed to reduce the number of variables to analyse.

Note 2: Average monthly precipitation per region (left) and inter-annual variability of precipitation per region expressed as standard deviation (sd). The black dotted line indicates the average rainfall and standard deviation over all regions.

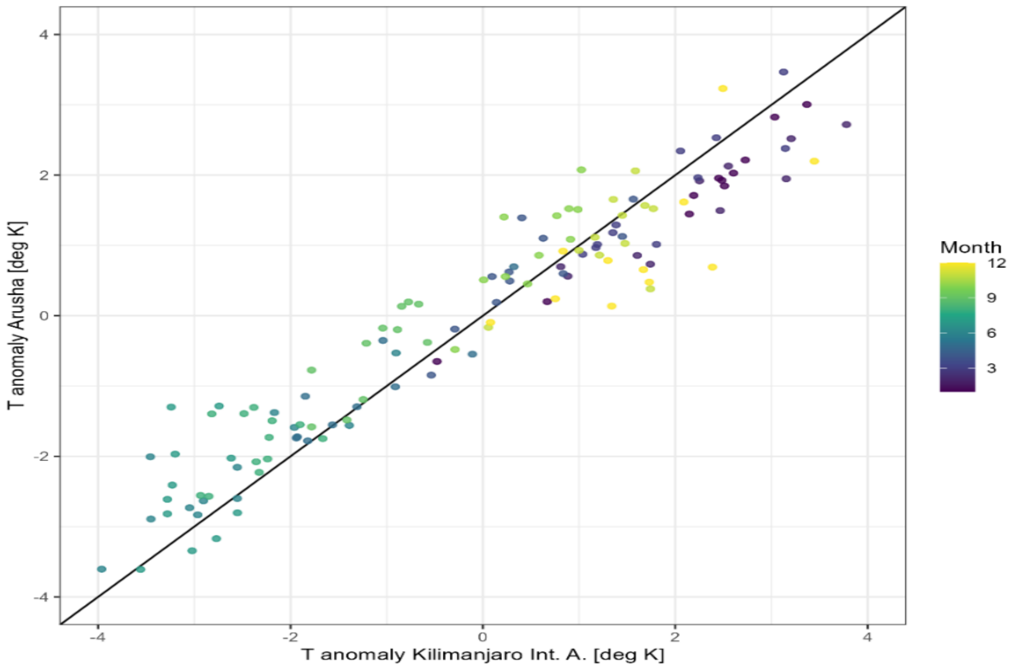


**Supplementary 4.** Monthly temperatures and precipitation

Note 1: Figure shows the comparison of the monthly temperature deviations from the norm from the weather station at Kilimanjaro International Airport and the weather station in Arusha (approximately 70 km south-west of the hospital). While the absolute temperatures of the two stations differ, the temperature anomalies are highly correlated. It was assumed that the temperature measurements at Kilimanjaro International Airport can be used as a proxy for the temperature anomalies experienced during pregnancies in north-east Tanzania.

Note 2: Average monthly anomaly of air temperature measured at the Kilimanjaro International Airport and in the city of Arusha. Although absolute temperatures between the stations vary, the regional anomaly is fairly consistent.

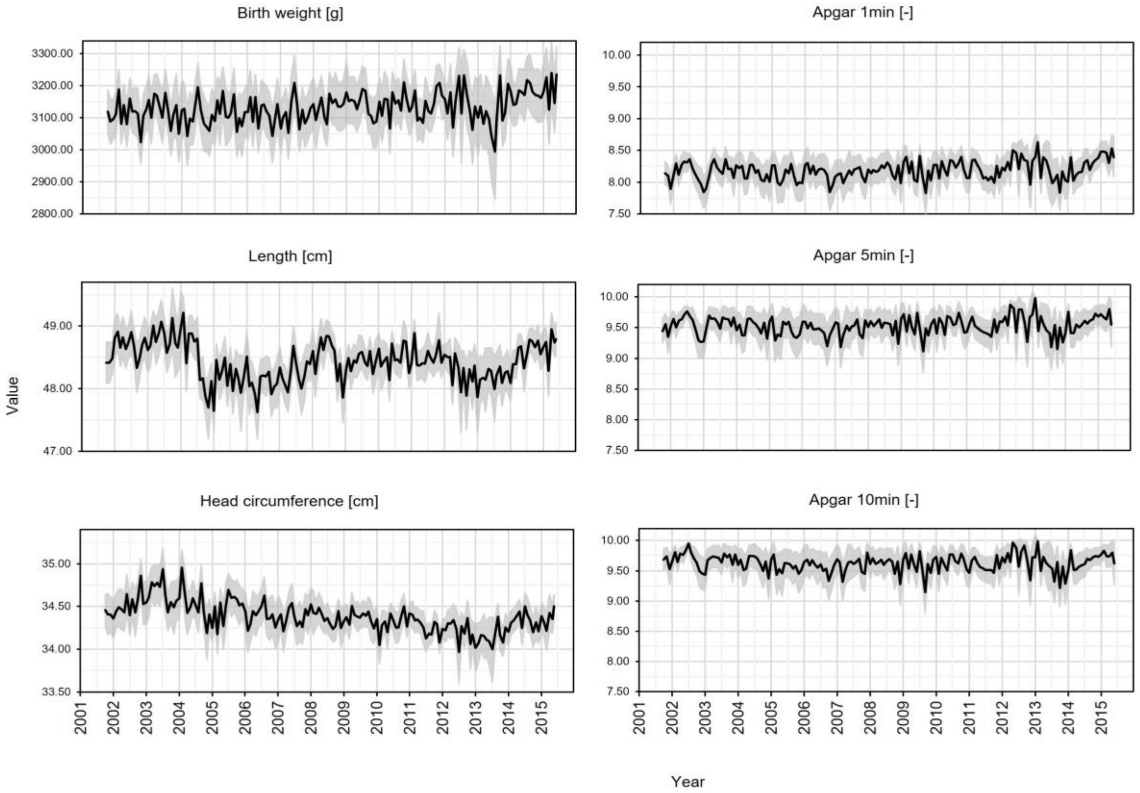


**Supplementary Figure 5**. Development of birth weight, neonatal length and neonatal head circumference from 2001-2015

Note: Grey shades represent monthly ranges from each outcome.

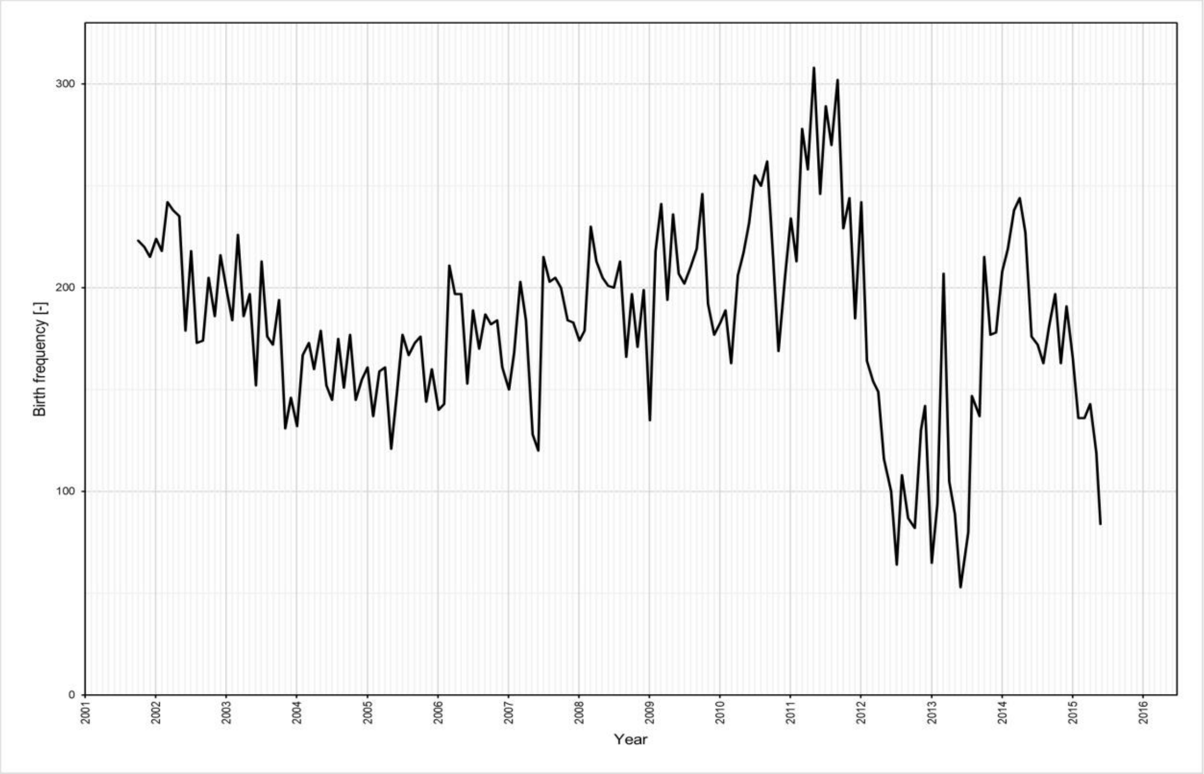


**Supplementary Figure 6.** Birth Frequency from 2001-2015

**Supplementary Table 1** Spearman correlations between weather conditions and birth outcomes.

| Variable | Birthweight | Length | Head circumference | Apgar at 1 min | Apgar at 5 min | Apgar at 10 min |
|---|---|---|---|---|---|---|
| NcDA 30°C | | | | | | |
| Preconception | -.001 | -.012* | -.034** | -.006 | .002 | -.008 |
| 1st trimester | .002 | -.003 | -.013* | -.006 | -.005 | -.009 |
| 2nd trimester | .012 | -.015 | .008 | .027** | .017* | .013 |
| 3rd trimester | .006 | -.004 | .006 | .013* | .014* | .011 |
| NdA 30°C | | | | | | |
| Preconception | -.005 | -.025* | -.024* | -.015* | -.012* | -.015 |
| 1st trimester | .003 | -.005 | .004 | -.004 | -.007 | -.008 |
| 2nd trimester | .011 | .016 | .015 | .022* | .016 | .012 |
| 3rd trimester | .001 | -.005 | -.015* | .008 | .013 | .007 |
| Rainfall | | | | | | |
| Preconception | -.005 | -.010 | .021** | .001 | .010 | .008 |
| 1st trimester | .000 | -.008 | .022** | .006 | .008 | .001 |
| 2nd trimester | .005 | -.001 | -.005 | .001 | -.001 | .003 |
| 3rd trimester | -.010 | -.015* | -.007 | -.017* | -.013* | -.004 |
| EDI | | | | | | |
| Preconception | -.005 | -.018** | -.004 | .005 | .012* | .006 |
| 1st trimester | .005 | -.008 | -.003 | .001 | .005 | -.003 |
| 2nd trimester | .002 | -.008 | -.025*** | .008 | .006 | .006 |
| 3rd trimester | .001 | -.014* | .022*** | -.007 | -.007 | -.003 |
| MEI | | | | | | |
| Preconception | -.008 | .013* | .068** | .008 | .002 | .001 |
| 1st trimester | -.003 | .014* | .048** | .009 | .008 | .004 |
| 2nd trimester | -.011 | .004 | .043** | -.003 | -.007 | -.008 |
| 3rd trimester | -.005 | .011 | .038** | -.007 | -.012* | -.009 |
| Harvest | .013* | .002 | -.002 | .019* | .009 | .011 |
